# Genome-wide Discovery for Diabetes-Dependent Triglycerides-Associated Loci

**DOI:** 10.1101/2022.01.06.22268848

**Authors:** Margaret Sunitha Selvaraj, Kaavya Paruchuri, Sara Haidermota, Rachel Bernardo, Stephen S. Rich, Gina M. Peloso, Pradeep Natarajan

## Abstract

We aimed to discover loci associated with triglyceride (TG) levels in the context of type 2 diabetes (T2D). We conducted a genome-wide association study (GWAS) in 424,120 genotyped participants of the UK Biobank (UKB) with T2D status and TG levels. We stratified the cohort based on T2D status and conducted association analyses of TG levels for genetic variants with minor allele count (MAC) at least 20 in each stratum. Effect differences of genetic variants by T2D status were determined by Cochran’s Q-test and we validated the significantly associated variants in the Mass General Brigham Biobank (MGBB). Among 21,176 T2D and 402,944 non-T2D samples from UKB, stratified GWAS identified 19 and 315 genomic risk loci significantly associated with TG levels, respectively. Only chr6p21.32 exhibited genome-wide significant heterogeneity (I^2^=98.4%; p_heterogeneity_=2.1×10^−15^), with log(TG) effect estimates of -0.066 (95%CI: - 0.082, -0.050) and 0.002 (95%CI: -0.002, 0.006) for T2D and non-T2D, respectively. The lead variant rs9274619:A (allele frequency 0.095) is located 2Kb upstream of the *HLA-DQB1* gene. We replicated this finding among 25,137 participants (6,951 T2D cases) of MGBB (p_heterogeneity_=9.5×10^−3^). Phenome-wide interaction association analyses showed that the lead variant was strongly associated with a concomitant diagnosis of type 1 diabetes (T1D) as well as diabetes-associated complications. In conclusion, we identified an intergenic variant near *HLA-DQB1* significantly associates with decreased triglycerides only among those with T2D and highlights an immune overlap with T1D.

## Introduction

Diabetes, largely due to type 2 diabetes (T2D), was estimated to afflict 9.3% of population in 2019 and projected to increase to 10.9% by 2045 (1). Despite ongoing scientific advances (2), T2D remains a leading cause of morbidity and mortality in the US and increasingly worldwide (3) (4). Novel approaches to discover the factors influencing T2D-related metabolic alterations may yield new insights toward the prevention of T2D-related complications.

Plasma lipid, particularly triglycerides (TG), alterations represent early metabolic changes linked to insulin resistance. Hypertriglyceridemia is often observed among individuals at risk for T2D and is more severe among individuals with poorly controlled T2D (5). Enhanced hepatic secretion of TG rich lipoproteins due to insulin resistance and delayed clearance involving lipoprotein lipase-mediated lipolysis may further exacerbate hypertriglyceridemia (6). Hypertriglyceridemia is an independent predictor of cardiovascular disease in T2D (7) (8), as well as a predictor of T2D itself (9). Characterizing the genetic determinants of TG concentrations specific to those with T2D may yield new insights into diabetes pathogenesis and complications.

Here, we tested the hypothesis that there are genetic variants associated with TG levels specific to T2D using GWAS and heterogeneity analysis in 424,120 participants of the UKB. Further, we assessed the role of the identified lead variant for multiple diabetes-related phenotypes.

## Results

### Baseline characteristics

The overall study schematic is depicted in **Supplementary Fig. 1**. Among 424,120 and 25,137 included samples, 21,176 (5.0%) and 6,951 (27.7%) of samples had T2D in UKB and MGBB, respectively. Overall UKB was composed of participants with a mean age (standard deviation [SD]) of 56.6 (8.1) years, 195,966 (46.2%) male, and 356,023 (83.9%) White British self-reported race. MGBB participants were mean 62.1 (16.2) years, 11,579 (46.1%) male, and 21,172 (84.2%) White British self-reported race. As expected, individuals with T2D versus non-T2D had greater median TG concentrations in both cohorts (**Table 1**).

**Table 1:** Baseline characteristics for discovery and replication cohorts: Distribution of samples across the T2D strata in discovery (UKB) and replication (MGBB) cohorts are provided. Number of samples by gender, ancestry and lipid lowering medications are documented. Lipid measurements for the four main lipid class is tabulated based on T2D strata. HLD-C – High-Density Lipoprotein Cholesterol; IQR – Inter quartile range; LDL-C – Low-Density Lipoprotein Cholesterol; MGBB – Mass General Brigham Biobank; SD – Standard Deviation; T2D – Type 2 Diabetes; TG – Triglycerides; TC – Total Cholesterol; UKB – UK Biobank.

| Cohorts | UK Biobank |  | Mass General Brigham Biobank |  |
| --- | --- | --- | --- | --- |
| Strata | T2D | non-T2D | T2D | non-T2D |
| Number of samples (%) | 21176(4.99) | 402944(95.01) | 6951(27.65) | 18185(72.34) |
| Age (years) mean (SD) | 60.22(6.87) | 56.36(8.11) | 67.79(13.28) | 59.93(16.73) |
| Female samples (%) | 8107(38.28) | 220047(54.60) | 3283(47.23) | 10274(56.40) |
| European samples (%) | 16636(78.56) | 339387(84.23) | 5457(78.51) | 15715(86.42) |
| Lipid lowering medication prescription (%) | 13433(63.44) | 56045(13.91) | 2197(31.61) | 2625(14.43) |
| TG concentration (mg/dL) median [IQR] | 171.83[125.09] | 129.58[95.66] | 125.00[93.00] | 97.00[70.00] |
| HDL-C concentration (mg/dL) mean (SD) | 46.04(12.44) | 56.51(14.71) | 49.78(17.82) | 58.88(19.80) |
| LDL-C concentration (mg/dL) mean (SD) | 111.59(34.17) | 138.98(33.04) | 90.74(35.88) | 104.08(35.89) |
| TC concentration (mg/dL) mean (SD) | 183.09(45.98) | 222.06(43.27) | 168.91(44.12) | 185.39(42.83) |

### T2D-stratified GWAS of TG identified an associated locus on chromosome 6

We performed GWAS on normalized natural log TG stratified by T2D status in the discovery cohort. Among the 402,944 non-T2D samples, 315 significant loci were identified. Among the 21,176 T2D samples, 19 significant loci were identified (**Supplementary Fig. 2, Supplementary Table 1**). We then assessed for differential TG effects for 67M variants by T2D status using Cochran’s Q-test for heterogeneity. We identified 478 variants which were genome-wide significant, all at chr6p21.32 (lead variant: rs9274619:G>A; I^2^=98.4%; p_heterogeneity_=2×10^−15^) (**Fig. 1**). The most significantly heterogenous variant was an intergenic variant near the *HLA-DQB1* gene (**Supplementary Fig. 3**), where the minor allele (frequency 0.095) decreases natural log TG among those with T2D but yields no difference among those with non-T2D (T2D group: beta=-0.066, p-value=3.9×10^−15^; non-T2D group: beta=0.002, p-value=0.21; p_interaction_=1.9×10^−11^). We observed that the difference test (Z_Diff_) identified this top significant lead variant (rs9274619:A; Z_diff_= -7.935) in the HLA locus as well. Since BMI is associated with both TG and T2D, we used scaled BMI as an additional covariate in testing the association of the identified lead variant with TG. We observed evidence for persistent albeit attenuated interaction between rs9274619:A and T2D after adjusting for BMI (p_interaction_=2.9×10^−7^). We observed a slight reduction in effects in the interaction model with T2D (beta_interaction_=-0.046), which shows that BMI has a confounding effect.

**Figure 1:**
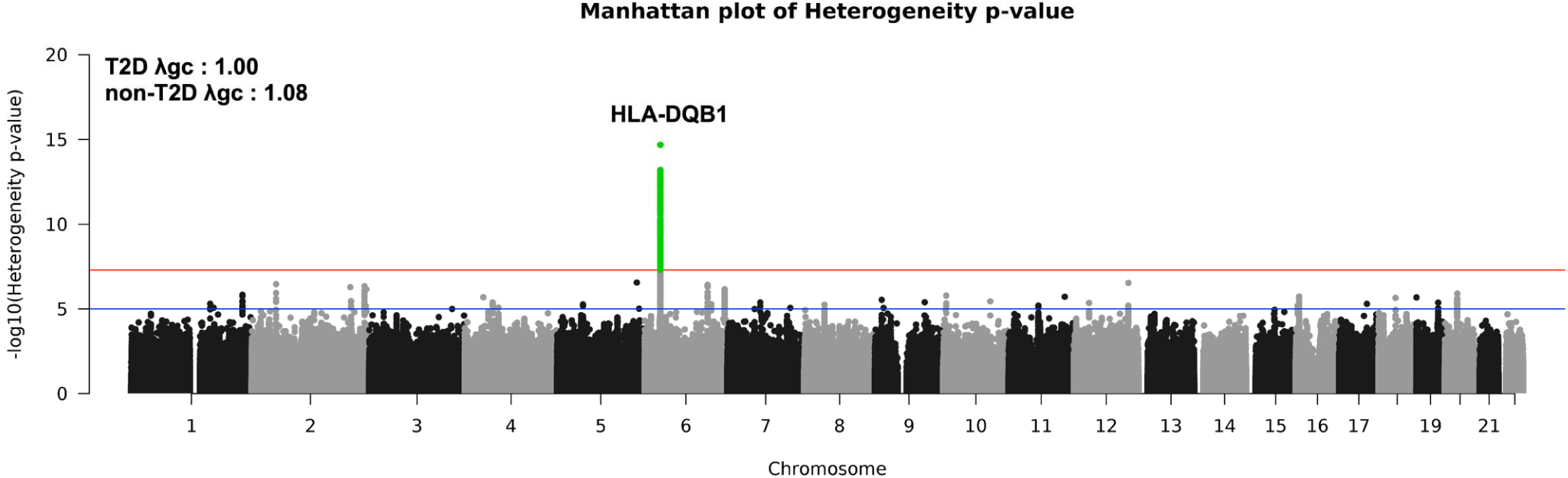
Genome-wide heterogeneity between T2D strata. Manhattan plot for heterogeneity p-values comparing T2D and non-T2D groups. Only one locus achieved genome-wide significance and the corresponding variants are colored in green. Lambda GC values from GWAS stratified by T2D status is shown in the figure. Red line: Genome-wide significance (p-value=5×10^−8^), Blue line: Suggestive significance (p-value=1×10^−5^). GWAS – Genome wide association studies; T2D – Type 2 Diabetes.

Individuals with T2D with lower TG concentrations were enriched for rs9274619:A (**Fig. 2A**). Among individuals with normal TG (i.e., <150 mg/dL), rs9274619:A was associated with T2D by 1.23-fold (95% CI 1.15,1.29; p-value 2.8×10^−11^). However, among individuals with TG > 450 mg/dl, rs9274619:A was not associated with T2D (OR 0.96, 95% CI 0.76-1.18; p-value 0.67). We replicated the findings in an independent cohort of 25,137 participants (6,951 T2D cases) of MGBB (p_heterogeneity_=9.5×10^−3^) (**Fig. 2B**). Additionally, we evaluated the association of the lead variant interacting with T2D status with other lipids in discovery cohort (**Supplementary Table 2**). We observed a significant interaction between rs9274619:A and T2D on HDL-C (p_interaction_=2.9×10^−8^) with higher concentrations among those with T2D, and nominally greater reductions in LDL-C among those with T2D (p_interaction_=6.0×10^−4^).

**Figure 2:**
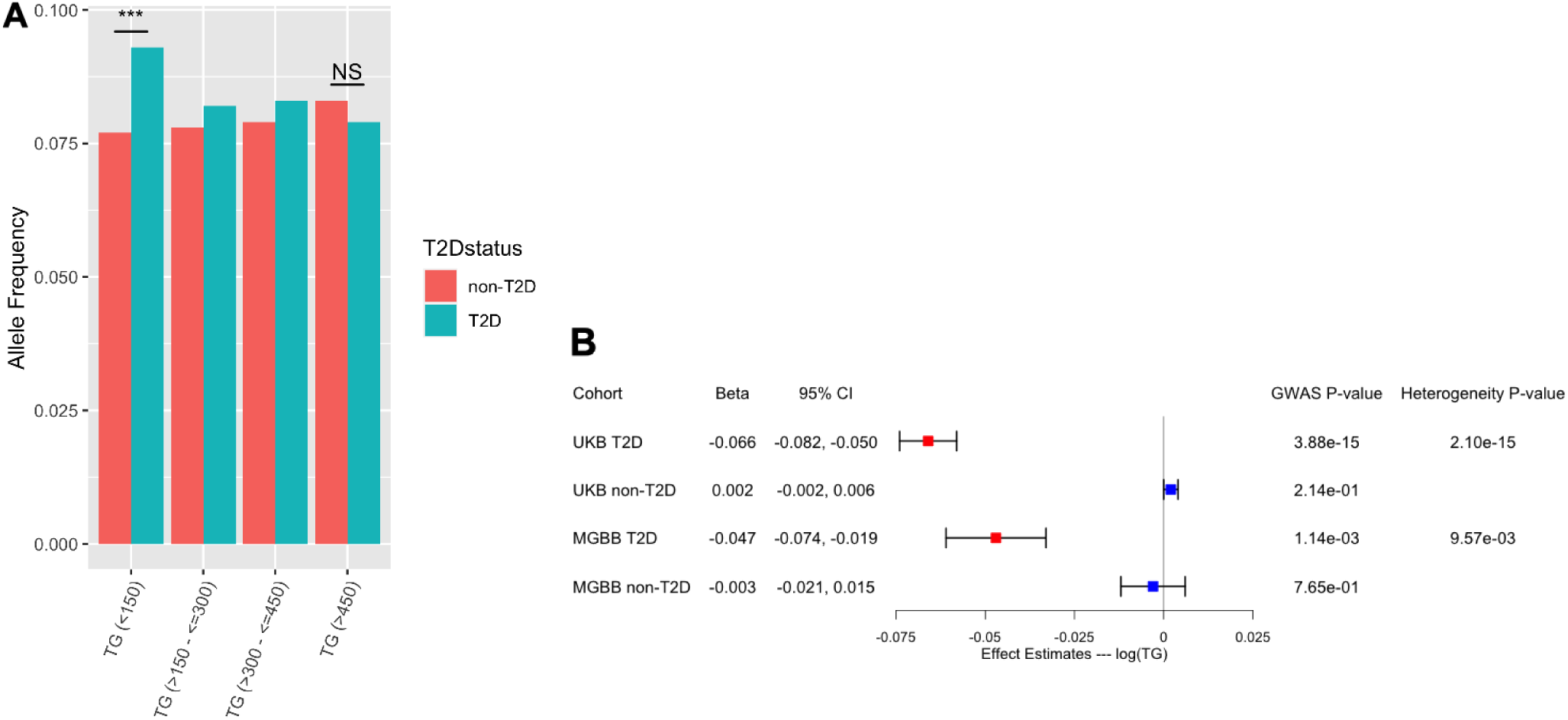
HLA-DQB1 rs9274619:A significantly interacts with T2D on triglycerides. A) Allele frequency of rs9274619:A in T2D and non-T2D samples grouped by raw TG values. Significance of samples proportions between the groups was assessed using Fisher’s exact test for the lower and higher TG bins. B) GWAS and heterogeneity statistics of the lead variant rs9274619:A at the *HLA-DQB1* locus from discovery (UKB) and replication (MGBB) cohorts based on T2D stratification. CI – Confidence Intervals; GWAS – Genome wide association studies; HLA – Human Leukocyte Antigens; MGBB – Mass General Brigham Biobank; NS – Non Significant; T2D – Type 2 Diabetes; TG – Triglycerides; UKB – UK Biobank.

We next bioinformatically prioritized the putative causal gene responsible for the T2D-dependent TG genetic association observed. Using T2D GWAS summary statistics PoPS prioritized the *HLA-DQB1* gene to be one of the top 20 genes along with other known TG genes such as *APOE, LPL* and *APOB*. However, *HLA-DQB1* was not prioritized in the non-T2D GWAS (**Supplementary Table 3**). Intersecting rs9274619:A with GTEx eQTL data for five different tissues (**Methods**) shows that the variant is an eQTL for multiple HLA genes including *HLA-DQB1* but more significantly for *HLA-DQA2* and *HLA-DRB6* (**Supplementary Table 4**). We curated all the eQTLs of the three HLA genes from GTEx database, T2D GWAS and correlated the Z-scores. eQTLs of all three HLA genes had a similar degree of correlations, but with opposite directions (**Supplementary Fig. 4**). We further interrogated pQTL and mQTL data. rs9274619:A is a pQTL for *HLA-DQA2* (beta=0.31; p-value=6.2×10^−14^) but it is an mQTL for multiple CpG regions at genome-wide significance. We identified 133 *cis*-associations and mapped the CpGs to Illumina HumanMethylation450 BeadChip (Illumina Inc., San Diego, USA) identification numbers (GEO data: GPL13534) to obtain the corresponding genes. Multiple HLA genes and other genes in chromosome 6 were identified (**Supplementary Table 5**). Gene prioritization using PoPS and QTL curation identified multiple HLA-genes (**Supplementary Fig. 5**).

### rs9274619:A tags HLA-DQB1*0302

Since the significant locus was at the HLA region, we correlated the rs9274619:A with 362 imputed HLA genotypes from 11 classes in the UKB. DQB1 and DQA1 were the most strongly correlated with rs9274619:A. Furthermore, DQB1_302 and DQA1_301 were most strongly correlated with rs9274619:A (DQB1_302: r=0.95, p_correlation_<3.83×10^−313^; DQA1_301: r=0.62, p_correlation_<3.83×10^−313^) (**Supplementary Fig. 6A**). We subsequently tested the interaction of all 362 HLA genotypes with T2D status on log(TG) as outcome. From this focused assessment of 362 HLA genotypes (**Supplementary Table 6**), 7 passed Bonferroni corrected significance (0.05/362=1×10^−4^) (**Supplementary Fig. 6B**). Consistent with our discovery and correlation analyses, only DQB1_302 had a genome-wide significant interaction (p_interaction_=1.05×10^−9^).

Although rs9274619:A is associated with increased expression of *HLA-DQA2* gene in eQTL and pQTL analysis, alleles from these HLA types were not previously imputed in UKB.

### Phenome-wide interaction analyses implicates multiple diabetes-related complications

We assessed the interactions between the rs9274619:A and T2D with 1567 disease conditions as outcomes (combination of incidence and prevalence) adjusted for all the covariates (age, age^2^, sex, race, PC1-10). Using a Bonferroni correction (0.05/1567=3.19×10^−5^), 45 disease phenotypes exhibited significant interactions. The strongest interaction was for the concomitant diagnosis of type 1 diabetes (T1D) among those with T2D (p_interaction_<1.72×10^−274^) (**Supplementary Table 7**). We applied logistic regression models stratified by T2D status on the 45 significant phenotypes, while adjusting for all covariates as mentioned above (**Supplementary Table 8**). Multiple diabetes-related microvascular and macrovascular complications including hypoglycemia, retinopathy, polyneuropathy, angiopathy, atherosclerosis and osteomyelitis were significantly associated, with the T2D-specific TG-lowering rs9274619:A allele leading to increased risks (**Fig. 3**). However, this allele was associated with reduced odds for obesity and related phenotypes.

**Figure 3:**
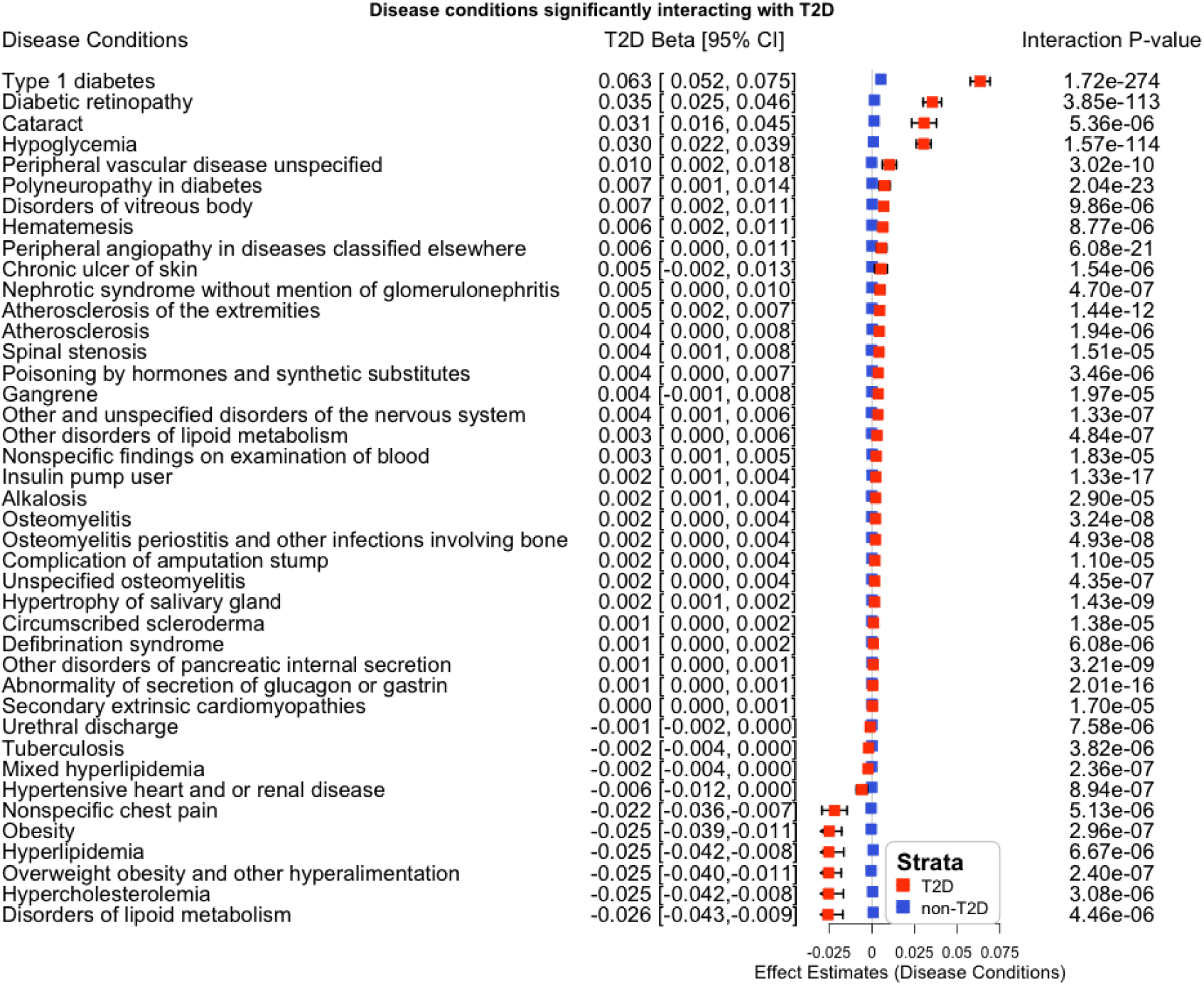
Phenome wide association (PheWAS) of disease conditions stratified by T2D status. Interaction between the rs9274619:A and T2D with multiple disease conditions as outcomes (combination of incidence and prevalence) was modeled while adjusting for all covariates (sex, age, age^2^, race, PC1-10). Bonferroni corrected 45 significant phenotypes were stratified by T2D status and analyzed using logistic regression. The T2D effect estimates for rs9274619:A and the interaction p-value are documented, the disease conditions are ordered based on T2D beta. From the 45 disease conditions tested, highly correlated Type1 diabetic conditions were removed while plotting the figure. PC – Principal Components; PheWAS – Phenome Wide Association Studies; T2D – Type 2 Diabetes.

Since HLA-DQB1 is a GWAS locus for an overlap between T1D and T2D previously referred to as latent autoimmune diabetes in adults (LADA) (10), as well as T1D itself (11), we further explored the relationships between the rs9274619:A, T1D, T2D, and their respective interactions on TGs (**Supplementary Table 9**). The TG-lowering rs9274619:A was strongly associated with T1D after adjusting for T2D (p-value=8.6×10^−113^), but not significantly associated with T2D status after adjusting for T1D (p-value=0.29). However, when assessing for interactions on TGs, there was still a significant interaction with T2D independent of T1D (beta_interaction_=-0.055; p_interaction_=5.28×10^−9^) and more strongly with T1D independently of T2D (beta_interaction_=-0.252; p_interaction_=5.53×10^−49^). Furthermore, we removed T1D samples and the interaction of rs9274619:A with T2D on TG was nominally significant (beta_interaction_=-0.022; p_interaction_=2.9×10^−2^).

Cousminer *et al* reported four loci to be significantly associated with LADA (12), therefore we assessed T2D/TG interactions for these lead variants (**Supplementary Table 10**). None of the variants tested had a genome-wide significant interaction. The variant rs9273368 from *HLA-DQB1* (p_interaction_=2.58×10^−5^) was genome-wide significant in our heterogeneity analysis (p_heterogeneity_=1.8×10^−8^) and was in moderate LD with rs9274619:A (R^2^=0.3). The four LADA loci were examined for interactions for additional diabetes-related phenotypes as noted in **Supplementary Table 11**.

### Metabolic characterizations of interactions with T2D

We secondarily explored the relationship between the rs9274619:A, interacting with T2D, and relationships with other metabolic features in UKB. Both the main effects and interaction models were adjusted for all the covariates (age, age^2^, sex, race, PC1-10). Outcomes assessed included waist/hip ratio (WHR), body mass index (BMI), macronutrients from 24-hour dietary recall surveys, and 60 plasma biomarkers (**Supplementary Table 12-13**). Several features were associated with the rs9274619:A itself, including increased eosinophil and neutrophil counts as well as hemoglobin A1c and C-reactive protein concentrations. We identified several outcomes that demonstrated differential association by T2D status in interaction testing (alpha 0.05/72=6.94×10^−4^). The TG-lowering rs9274619:A allele was associated with greater concentrations for T2D vs non-T2D for hemoglobin A1c, sex hormone binding globulin, HDL-C, glucose, and apolipoprotein-A1 concentrations. However, the TG-lowering rs9274619:A allele was associated with reduced concentrations for T2D vs non-T2D for urate, BMI, WHR, and reticulocyte count (**Supplementary Table 12-13**).

We further assessed lipid metabolomic data comprising 102,575 UKB samples (5200 T2D cases) and 249 metabolites for interactions. For each normalized metabolomic phenotype we analyzed the main effect of rs9274619:A and the interaction of rs9274619:A with T2D, with both models adjusted for all aforementioned covariates. Using a Bonferroni correction (0.05/249=2.01×10^−4^), we identified 6 metabolomic features associated with the rs9274619:A and only 1 interaction with T2D (**Supplementary Table 14-15**). With respect to the interaction detected, we observed that the average diameter for LDL particles among rs9274619:A carriers was greater for T2D versus non-T2D.

## Discussion

Independent GWAS studies have identified multiple loci strongly associated with TG and T2D separately (13),(14), and we now observe a variant tagging HLA-DQ1B*0302 associated with TGs only among those with T2D and not among those without T2D. Despite being associated with reduced TGs specifically in T2D, the lead variant is associated with greater diabetes-related complications and an overlap with T1D.

Our study permits several conclusions regarding T2D pathogenesis. First, our study highlights heterogeneous metabolism among individuals with adult-onset diabetes. Hyperinsulinemia contributes to reduced hydrolysis and clearance of TG rich lipoproteins and thus persistence of these atherogenic lipoproteins toward heightened macrovascular risk (15). TGs have more moderate associations with microvascular complications among diabetics (16). Indeed, glycemic control is a more potent risk factor for microvascular complications. Here, we find that an immune-related locus (i.e., *HLA-DQB1* genotype of major histocompatibility class II) linked to reduced TGs interestingly associates with greater microvascular versus macrovascular risk among individuals with T2D. Lipid homeostasis plays a key role in immune cells, where lipids are key constituents of major histocompatibility complex molecules and other cell membrane microdomains (17). These observations highlight complementary roles of immune dysfunction and hyperinsulinemia in adult-onset diabetes pathogenesis.

Second, the distinct lipid pattern observed by HLA-DQB1*0302 genotype may reflect etiologically distinct subgroups of adult-onset diabetes. The HLA-DQB1*0302 has long been recognized as a very strong risk factor for T1D and tags the potent T1D DR4 risk haplotype (18),(19),(20),(21). Approximately 2-12% of adults diagnosed with type 2 diabetes have glutamic acid decarboxylase autoantibodies (GADA), thereby leading to the proposed term of latent autoimmune diabetes in adults (LADA) (22),(23),(24),(25). Such individuals are often classified as T2D because they typically do not initially require insulin. Indeed, a recent GWAS of LADA showed that *HLA-DQB1* was the most significant locus(12). Recent data-driven approaches to cluster diabetes have grouped diabetes with GADA, traditionally classified as T1D or LADA, as severe autoimmune diabetes (SAID) (26). Consistent with separate T1D and LADA analyses, the *HLA-DQB1* locus is significantly associated with SAID unlike with other diabetes subgroups (27). Thus, the relatively reduced TG concentrations among adults classified as having T2D and the *HLA-DQB1* risk allele may reflect the lack of hypertriglyceridemia typically observed with more typical hyperinsulinemic T2D.

Third, TG concentrations among individuals diagnosed with T2D may help identify individuals with features more consistent with T1D. GADA testing only in adult-onset diabetics with normal TGs would optimize diagnostic yield. Furthermore, with increasingly available genotyping through expanding research testing and widely used direct-to-consumer approaches, HLA genotypes may further improve efficiency of testing. While large-scale randomized controlled trials for LADA are lacking, expert consensus recommend personalized management approaches deviating from conventional T2D and surveillance management (28). Such approaches include biomarker-based surveillance of residual beta cell function and to determine insulin initiation.

A few limitations of our study deserve mention. First, causal gene prioritization through multiple methods did not converge on a single gene. Whether our observations reflect coordinated regulation merits further study. Based on the current results we were not able to elucidate the exact mechanism of TG lowering by HLA rs9274619:A in the context of T2D. Second, we found that after adjusting for T1D, the association between T2D and rs9274619:A was no longer significant, indicating that part of the association between TG and rs9274619:A may be due to a concomitant diagnosis of T1D. Third, UKB and MGBB are predominantly White and this may limit generalizability to other genetic backgrounds. Finally, Cochran’s Q test is under powered to detect differences in heterogeneity of effect sizes and we may have missed other loci with differences in effects. However, we also performed a difference test and found similar results to using the Cochran’s Q test.

In conclusion, we observed that HLA-DQB1*0302 is associated with reduced TGs only among adults with diabetes. Presence of this allele reflects an autoinflammatory subgroup of adult-onset diabetes most consistent with T1D, without characteristic hyperinsulinemia and thus relatively reduced TG concentrations. Among individuals classified as T2D, these individuals have greater risks for diabetes-associated complications.

## Methods

### Study Participants

We used the UK Biobank (UKB), which is a prospective population-based cohort composed of approximately 500,000 samples with rich phenotypic and genotypic information, as the discovery cohort (29). UKB includes volunteer residents of the UK aged 40 to 69 years recruited during 2006-2010. The phenotypic information includes details on lifestyle, medical history, food habits, weight, height, body measurements, scans, blood routines, and electronic medical record (EMR) coded data. Out of 488,377 total individuals, we removed unconsented individuals and samples with >10% genotypic missingness thereby retaining 424,120 individuals for whom the TGs were also available. All individuals provided informed consent per the UKB primary protocol. Secondary use of these data was approved by the Massachusetts General Hospital Institutional Review Board (protocol 2021P002228) and was facilitated through UKB application 7089.

We used data from the Mass General Brigham Biobank (MGBB), which comprises volunteer patients of the large Mass General Brigham Healthcare system in Massachusetts with greater than 105,000 participants, as replication (30). In total 36,424 randomly selected individuals were genotyped using three versions of the Multi-Ethnic Genotyping Array (MEGA) Single-Nucleotide Polymorphism (SNP) array (Multiethnic Exome Global (Meg), Human multi-ethnic array (Mega), Expanded multi-ethnic genotyping array (Megex)). Out of 36,424 individuals, we retained 25,137 samples for whom T2D status and TG measurements were available for the current study. All individuals provided informed consent per the MGBB primary protocol. Secondary use of these data was approved by the Massachusetts General Hospital Institutional Review Board (protocol 2020P000904).

### Phenotypes

In the UKB, we defined T2D based on self-reported status (data field 20002) and ICD10 codes E11:0-9 (data fields 41202, 41204, 40001, and 40002). The first instance of TG measurement (data field 30870) was defined as the primary lipid phenotype of interests. We also included other lipid levels as secondary outcomes: total cholesterol (TC) (data field 30690), low-density lipoprotein cholesterol (LDL-C) (data field 30780), and high-density lipoprotein cholesterol (HDL-C) (data field 30760). TG measurements were converted to mg/dL by multiplying mmol/L values by 88.57 and natural log transformed. TC, LDL-C and HDL-C values in mmol/L were converted to mg/dL by multiplying 38.67. When lipid-lowering medications were prescribed, TC measurements were divided by 0.8 and LDL-C by 0.7, as previously done (14). All four lipid measurements were further inverse rank normalized to the residuals scaled by the standard deviation, where the model was adjusted for covariates (sex, age, age^2^, PC1-10).

We curated multiple diseases for UKB samples into phecodes for PheWAS analysis. The PheWAS R package (version PheWAS_0.99.5-4) was used to map ICD codes to phecodes based on the phecode map 1.2 and 1.2b1 from https://phewascatalog.org/ (31). Codes that failed to map were excluded, which were relatively few and often procedural. Mapped codes were defined as multiple disease conditions and specified as incident or prevalent based on the time of sample collection. Next, we obtained the secondary phenotypes, which included waist circumference (data field 48), hip circumference (data field 49), body mass index (BMI) (data field 23104) and 24-hour diet recall (data field 110001) for downstream analysis. Additionally, we included blood biochemistry (category id 18518) and count (category id 9081) measures. These phenotypes were normalized to a mean 0 and standard deviation 1 for analysis. We obtained the NMR metabolic biomarkers generated by Nightingale Health (Helsinki, Finland) from the first tranche of 249 metabolic biomarkers in 118,032 UKB participants (32). We included 102,528 samples that intersected with the discovery cohort and each of the metabolites were inverse rank normalized and regressed against the covariates (age, age^2^, sex, race, PC1-10). The residuals were used as our phenotypes in analyses.

In MGBB, electronic health record (EHR) data were used to define incident and prevalent cases based on enrollment date and ICD-9/ICD-10 codes on clinical phenotype definitions from phecode groups(33) (34), where samples with phecode 250.2X were defined as T2D in our study. Similarly, lipid test results, medication information, demographic status of genotyped samples were curated from EHR records. LDL-C was measured directly or calculated using Friedewald equation when TG were <400 mg/dL, all lipid measurements were in mg/dL units. The lipid measurements closest to sequencing date was curated. A sample was defined as on statin medication, if statin treatment was prescribed within the last one year of the sequencing date. We performed phenotype harmonization and normalization for the validation data as described above.

### Genotypes

Genetic data from 488,377 UKB samples were assayed using two similar genotyping arrays from Affymetrix (Santa Clara, CA): i) Applied Biosystems UK BiLEVE Axiom Array ii) Applied Biosystems UK Biobank Axiom Array. 49,950 participants with 807,411 markers were genotyped at using the Applied Biosystems UK BiLEVE Axiom Array and 438,427 participants with 825,927 markers were genotyped using the closely related Applied Biosystems UK Biobank Axiom Array. Both arrays shared 95% of marker content and the UK Biobank Axiom array was chosen to capture genome-wide genetic variation (single nucleotide polymorphism (SNPs) and short insertions and deletions (indels)) (29). The imputation from the UKB array-derived genotypes was performed using merged UK10K and 1000 Genomes phase 3 reference panels (35) and was combined to the Haplotype Reference Consortium (HRC) (36) panel using IMPUTE4 program (https://jmarchini.org/software/) as implemented in IMPUTE2 (37). We obtained the UKB imputed human leukocyte antigen (HLA) genotypes (data field 22182) composed of classical allelic variation of 11 HLA types (A, B, C, DRB5, DRB4, DRB3, DRB1, DQB1, DQA1, DPB1, DPA1). HLA imputation from allele pairs was performed using HLA*IMP:02 in the UKB, as previously described (29). Genotypic data in MGBB cohort was generated using three different arrays (Multiethnic Exome Global [MEG], Human multi-ethnic array [MEGA], Expanded multi-ethnic genotyping array [MEGEX]) from Illumina (San Diego, CA).

### Statistical analysis

We performed genome wide association analysis (GWAS) stratified based on T2D status. We first performed quality control (QC) of the full UKB dataset regardless of T2D status by applying additional filters, including minor allele frequency (MAF) < 1%, Hardy-Weinberg equilibrium p-value not exceeding 1×10^−15^ and genotype missingness > 10% to filter variants, and sample-level genotype missingness > 10%. The QC-passed dataset was used to create NULL model with sex, age, age^2^, genotype array, race and PC1-10 as covariates. We employed REGENIE with leave-one-out-cross-validation (LOOCV) (38) approach adjusted for covariates stated above to perform GWAS on UKB imputed data with minor allele count (MAC) 20 in both T2D and non-T2D samples, independently. We annotated the genomic risk-loci from GWAS summary statistics using FUMA(39).

We tested for differences in effect estimates per genotype by T2D status using Cochran’s Q-test for heterogeneity in the METAL package (40). For the genome-wide heterogeneity assessment, we used the conventional alpha threshold of 5×10^−8^ to assign statistical significance accounting for multiple-hypothesis testing. We validated the outcomes from the Cochran’s Q-test using the Difference Test (41). Significant lead variant in the discovery dataset were replicated at an alpha threshold of 0.05 accounting for the single SNP assessed.

The significant and replicated variant (lead variant) was pursued for further downstream analysis. Given its genomic location, we correlated the UKB imputed classical HLA genotypes with the significant lead variant using corplot R-package (method-pearson; version-0.90). We performed regression-based interaction analyses using the model where adiposity-related, diet-related and other blood biomarker phenotypes were separately analyzed with the lead variant along with T2D status. The regression analysis (main and interaction model) was carried out in R, adjusting for sex, age, age^2^, genotype array, race, and PC1-10 as covariates. Bonferroni corrected alpha threshold of 0.05/number of tests was considered statistically significant for these analyses. Fisher’s exact test was performed to test the significance of sample proportions among group of samples.

We implemented the Polygenic Priority Score (PoPS) enrichment method (42) for gene prioritization with the GWAS summary statistics. PoPS integrates multiple public bulk and single-cell expression datasets, protein-protein interaction and pathway databases to implement enrichment analysis using MAGMA (43) based gene association scores to identify top list of genes functionally linked to the phenotype of interest. We complementarily performed quantitative trait locus (QTLs) interrogations using multiple publicly available datasets. We downloaded expression quantitative trait locus (eQTLs) data from GTEx (v8_eQTL_all_associations) database (https://gtexportal.org/home/datasets) and curated significant hits (p-value < 5×10^−8^) for the lead SNP from 5 different tissues relevant to diabetes, lipids, and inflammation (i.e., Liver, Adipose Subcutaneous, Adipose Visceral Omentum, Whole Blood, and Pancreas). We utilized protein quantitative trait loci (pQTL) data in blood from the INTERVAL study (44) and GoDMC database (45) for methylation quantitative trait loci (mQTL) in blood to curate pQTLs and mQTLs related to the lead variant (p-value < 5×10^−8^).

## Data Availability

All data produced in the present study are available upon reasonable request to the authors

## Acknowledgements

We thank all the participants from UKB and MGBB. P.N. is supported by grants from the National Institutes of Health (R01HL142711, R01HL148050, R01HL151283, R01HL127564, R01HL148565, R01HL151152, R01DK125782), Fondation Leducq (TNE-18CVD04), and Massachusetts General Hospital (Fireman Chair). GMP is supported by NIH grants R01HL127564 and R01HL142711.

## Disclosures

P.N. reports grants from Amgen, Apple, AstraZeneca, Boston Scientific, and Novartis, personal fees from Apple, AstraZeneca, Blackstone Life Sciences, Foresite Labs, Genentech / Roche, Novartis, and TenSixteen Bio, equity in geneXwell, and TenSixteen Bio, co-founder of TenSixteen Bio, and spousal employment at Vertex, all unrelated to the present work.

## Notes

### Author Declarations

Secondary use of the data was approved by the Massachusetts General Hospital Institutional Review Board (protocol 2021P002228) and was facilitated through UKB application 7089. Secondary use of the data was approved by the Massachusetts General Hospital Institutional Review Board (protocol 2020P000904).

## References

1. Saeedi P, Petersohn I, Salpea P, Malanda B, Karuranga S, Unwin N, et al. Global and regional diabetes prevalence estimates for 2019 and projections for 2030 and 2045: Results from the International Diabetes Federation Diabetes Atlas, 9th edition. Diabetes Res Clin Pract. 2019 Nov;157:107843.

2. Gillies CL, Abrams KR, Lambert PC, Cooper NJ, Sutton AJ, Hsu RT, et al. Pharmacological and lifestyle interventions to prevent or delay type 2 diabetes in people with impaired glucose tolerance: systematic review and meta-analysis. BMJ. 2007 Feb 10;334(7588):299.

3. Tancredi M, Rosengren A, Svensson A-M, Kosiborod M, Pivodic A, Gudbjörnsdottir S, et al. Excess Mortality among Persons with Type 2 Diabetes. N Engl J Med. 2015 Oct 29;373(18):1720–32.

4. Wright AK, Kontopantelis E, Emsley R, Buchan I, Sattar N, Rutter MK, et al. Life Expectancy and Cause-Specific Mortality in Type 2 Diabetes: A Population-Based Cohort Study Quantifying Relationships in Ethnic Subgroups. Diabetes Care. 2017 Mar;40(3):338–45.

5. Low S, Khoo KCJ, Irwan B, Sum CF, Subramaniam T, Lim SC, et al. The role of triglyceride glucose index in development of Type 2 diabetes mellitus. Diabetes Res Clin Pract. 2018 Sep;143:43–9.

6. Pang J, Chan DC, Watts GF. Origin and therapy for hypertriglyceridaemia in type 2 diabetes. World J Diabetes. 2014 Apr 15;5(2):165–75.

7. Feher M, Greener M, Munro N. Persistent hypertriglyceridemia in statin-treated patients with type 2 diabetes mellitus. Diabetes Metab Syndr Obes Targets Ther. 2013;6:11–5.

8. Leiter LA, Lundman P, da Silva PM, Drexel H, Jünger C, Gitt AK, et al. Persistent lipid abnormalities in statin-treated patients with diabetes mellitus in Europe and Canada: results of the Dyslipidaemia International Study. Diabet Med J Br Diabet Assoc. 2011 Nov;28(11):1343–51.

9. Zhao J, Zhang Y, Wei F, Song J, Cao Z, Chen C, et al. Triglyceride is an independent predictor of type 2 diabetes among middle-aged and older adults: a prospective study with 8-year follow-ups in two cohorts. J Transl Med. 2019 Dec;17(1):403.

10. Basile KJ, Guy VC, Schwartz S, Grant SFA. Overlap of genetic susceptibility to type 1 diabetes, type 2 diabetes, and latent autoimmune diabetes in adults. Curr Diab Rep. 2014;14(11):550.

11. Noble JA, Valdes AM. Genetics of the HLA region in the prediction of type 1 diabetes. Curr Diab Rep. 2011 Dec;11(6):533–42.

12. Cousminer DL, Ahlqvist E, Mishra R, Andersen MK, Chesi A, Hawa MI, et al. First Genome-Wide Association Study of Latent Autoimmune Diabetes in Adults Reveals Novel Insights Linking Immune and Metabolic Diabetes. Diabetes Care. 2018 Nov;41(11):2396–403.

13. Scott RA, Scott LJ, Mägi R, Marullo L, Gaulton KJ, Kaakinen M, et al. An Expanded Genome-Wide Association Study of Type 2 Diabetes in Europeans. Diabetes. 2017 Nov;66(11):2888–902.

14. NHLBI TOPMed Lipids Working Group, Natarajan P, Peloso GM, Zekavat SM, Montasser M, Ganna A, et al. Deep-coverage whole genome sequences and blood lipids among 16,324 individuals. Nat Commun. 2018 Dec;9(1):3391.

15. Miller M, Stone NJ, Ballantyne C, Bittner V, Criqui MH, Ginsberg HN, et al. Triglycerides and Cardiovascular Disease: A Scientific Statement From the American Heart Association. Circulation. 2011 May 24;123(20):2292–333.

16. Sacks FM, Hermans MP, Fioretto P, Valensi P, Davis T, Horton E, et al. Association Between Plasma Triglycerides and High-Density Lipoprotein Cholesterol and Microvascular Kidney Disease and Retinopathy in Type 2 Diabetes Mellitus: A Global Case–Control Study in 13 Countries. Circulation. 2014 Mar 4;129(9):999–1008.

17. Hiltbold EM, Poloso NJ, Roche PA. MHC Class II-Peptide Complexes and APC Lipid Rafts Accumulate at the Immunological Synapse. J Immunol. 2003 Feb 1;170(3):1329–38.

18. Erlich H, Valdes AM, Noble J, Carlson JA, Varney M, Concannon P, et al. HLA DR-DQ Haplotypes and Genotypes and Type 1 Diabetes Risk: Analysis of the Type 1 Diabetes Genetics Consortium Families. Diabetes. 2008 Apr 1;57(4):1084–92.

19. Noble JA, Valdes AM, Varney MD, Carlson JA, Moonsamy P, Fear AL, et al. HLA Class I and Genetic Susceptibility to Type 1 Diabetes: Results From the Type 1 Diabetes Genetics Consortium. Diabetes. 2010 Nov 1;59(11):2972–9.

20. Noble JA, Erlich HA. Genetics of Type 1 Diabetes. Cold Spring Harb Perspect Med. 2012 Jan 1;2(1):a007732–a007732.

21. Jacobi T, Massier L, Klöting N, Horn K, Schuch A, Ahnert P, et al. HLA Class II Allele Analyses Implicate Common Genetic Components in Type 1 and Non– Insulin-Treated Type 2 Diabetes. J Clin Endocrinol Metab. 2020 Mar 1;105(3):e245–54.

22. Buzzetti R, Di Pietro S, Giaccari A, Petrone A, Locatelli M, Suraci C, et al. High Titer of Autoantibodies to GAD Identifies a Specific Phenotype of Adult-Onset Autoimmune Diabetes. Diabetes Care. 2007 Apr 1;30(4):932–8.

23. Radtke MA, Midthjell K, Nilsen TIL, Grill V. Heterogeneity of Patients With Latent Autoimmune Diabetes in Adults: Linkage to Autoimmunity Is Apparent Only in Those With Perceived Need for Insulin Treatment: Results from the Nord-Trondelag Health (HUNT) study. Diabetes Care. 2009 Feb 1;32(2):245–50.

24. Tuomi T, Santoro N, Caprio S, Cai M, Weng J, Groop L. The many faces of diabetes: a disease with increasing heterogeneity. The Lancet. 2014 Mar;383(9922):1084–94.

25. Turner R, Stratton I, Horton V, Manley S, Zimmet P, Mackay IR, et al. UKPDS 25: autoantibodies to islet-cell cytoplasm and glutamic acid decarboxylase for prediction of insulin requirement in type 2 diabetes. The Lancet. 1997 Nov;350(9087):1288–93.

26. Ahlqvist E, Storm P, Käräjämäki A, Martinell M, Dorkhan M, Carlsson A, et al. Novel subgroups of adult-onset diabetes and their association with outcomes: a data-driven cluster analysis of six variables. Lancet Diabetes Endocrinol. 2018 May;6(5):361–9.

27. Mansour Aly D, Dwivedi OP, Prasad RB, Käräjämäki A, Hjort R, Thangam M, et al. Genome-wide association analyses highlight etiological differences underlying newly defined subtypes of diabetes. Nat Genet. 2021 Nov;53(11):1534–42.

28. Buzzetti R, Tuomi T, Mauricio D, Pietropaolo M, Zhou Z, Pozzilli P, et al. Management of Latent Autoimmune Diabetes in Adults: A Consensus Statement From an International Expert Panel. Diabetes. 2020 Oct;69(10):2037–47.

29. Bycroft C, Freeman C, Petkova D, Band G, Elliott LT, Sharp K, et al. The UK Biobank resource with deep phenotyping and genomic data. Nature. 2018 Oct;562(7726):203–9.

30. Smoller J, Karlson E, Green R, Kathiresan S, MacArthur D, Talkowski M, et al. An eMERGE Clinical Center at Partners Personalized Medicine. J Pers Med. 2016 Jan 20;6(1):5.

31. Denny JC, Ritchie MD, Basford MA, Pulley JM, Bastarache L, Brown-Gentry K, et al. PheWAS: demonstrating the feasibility of a phenome-wide scan to discover gene-disease associations. Bioinformatics. 2010 May 1;26(9):1205–10.

32. Julkunen H, Cichońska A, Slagboom PE, Würtz P, Nightingale Health UK Biobank Initiative. Metabolic biomarker profiling for identification of susceptibility to severe pneumonia and COVID-19 in the general population. eLife. 2021 May 4;10:e63033.

33. Wei W-Q, Bastarache LA, Carroll RJ, Marlo JE, Osterman TJ, Gamazon ER, et al. Evaluating phecodes, clinical classification software, and ICD-9-CM codes for phenome-wide association studies in the electronic health record. PloS One. 2017;12(7):e0175508.

34. Wu P, Gifford A, Meng X, Li X, Campbell H, Varley T, et al. Mapping ICD-10 and ICD-10-CM Codes to Phecodes: Workflow Development and Initial Evaluation. JMIR Med Inform. 2019 Nov 29;7(4):e14325.

35. UK10K Consortium, Huang J, Howie B, McCarthy S, Memari Y, Walter K, et al. Improved imputation of low-frequency and rare variants using the UK10K haplotype reference panel. Nat Commun. 2015 Nov;6(1):8111.

36. the Haplotype Reference Consortium. A reference panel of 64,976 haplotypes for genotype imputation. Nat Genet. 2016 Oct;48(10):1279–83.

37. Howie B, Fuchsberger C, Stephens M, Marchini J, Abecasis GR. Fast and accurate genotype imputation in genome-wide association studies through pre-phasing. Nat Genet. 2012 Aug;44(8):955–9.

38. Mbatchou J, Barnard L, Backman J, Marcketta A, Kosmicki JA, Ziyatdinov A, et al. Computationally efficient whole-genome regression for quantitative and binary traits. Nat Genet. 2021 Jul;53(7):1097–103.

39. Watanabe K, Taskesen E, van Bochoven A, Posthuma D. Functional mapping and annotation of genetic associations with FUMA. Nat Commun. 2017 Dec;8(1):1826.

40. Willer CJ, Li Y, Abecasis GR. METAL: fast and efficient meta-analysis of genomewide association scans. Bioinformatics. 2010 Sep 1;26(17):2190–1.

41. Winkler TW, Justice AE, Cupples LA, Kronenberg F, Kutalik Z, Heid IM, et al. Approaches to detect genetic effects that differ between two strata in genome-wide meta-analyses: Recommendations based on a systematic evaluation. Meyre D, editor. PLOS ONE. 2017 Jul 27;12(7):e0181038.

42. Weeks EM, Ulirsch JC, Cheng NY, Trippe BL, Fine RS, Miao J, et al. Leveraging polygenic enrichments of gene features to predict genes underlying complex traits and diseases [Internet]. Genetic and Genomic Medicine; 2020 Sep [cited 2021 Sep 7]. Available from: http://medrxiv.org/lookup/doi/10.1101/2020.09.08.20190561

43. de Leeuw CA, Mooij JM, Heskes T, Posthuma D. MAGMA: Generalized Gene-Set Analysis of GWAS Data. Tang H, editor. PLOS Comput Biol. 2015 Apr 17;11(4):e1004219.

44. Sun BB, Maranville JC, Peters JE, Stacey D, Staley JR, Blackshaw J, et al. Genomic atlas of the human plasma proteome. Nature. 2018 Jun;558(7708):73–9.

45. Min JL, Hemani G, Hannon E, Dekkers KF, Castillo-Fernandez J, Luijk R, et al. Genomic and phenotypic insights from an atlas of genetic effects on DNA methylation. Nat Genet. 2021 Sep;53(9):1311–21.

