## Supplementary Text for "Genome-wide Discovery for Diabetes-Dependent Triglycerides-Associated Loci"

**Supplementary Table Legends**

**Supplementary Table 1:** Genomic risk loci identified from GWAS stratified by T2D status in UKB. 19 and 315 risk loci were identified by FUMA on T2D and non-T2D GWAS respectively. Significant loci and lead variants identified from FUMA analysis for each risk loci are documented.

GWAS – Genome wide association studies; UKB – UK Biobank; T2D – Type 2 Diabetes.

**Supplementary Table 2:** Summary statistics from linear regression model where rs9274619:A interacting with T2D is documented for log(TG) as outcome for the 3 lipids including LDL-C, HDL-C and TC. *HLA-DQB1* is enriched only in T2D and not in non-T2D.

HLD-C – High-Density Lipoprotein Cholesterol; LDL-C – Low-Density Lipoprotein Cholesterol; T2D – Type 2 Diabetes; TG – Triglycerides; TC – Total Cholesterol.

**Supplementary Table 3:** PoPS enrichment scores for the top100 genes from T2D and nonT2D GWAS summary statistics. The Genes are ordered based on PoPS scores within each strata.

T2D – Type 2 Diabetes.

**Supplementary Table 4:** eQTL – gene pairs for rs9274619:A was curated from GTEx data and genome significant hits were obtained (p-value=5x10^-8^) from 5 different tissues. Each associated gene is tabulated and ordered based on GTEx p-value.

eQTL – Expression Quantitative Trait Locus.

**Supplementary Table 5:** mQTL – CpG pairs for rs9274619:A and the corresponding genes that regulate the CpG regions was curated from GoDMC database where the genome significant hits were obtained (p-value=5x10^-8^) from the cis-associations. The data is ordered based on p-value and the corresponding CpG island regions were curated from Illumina HumanMethylation450 BeadChip resource.

mQTL –Methylation Quantitative Trait Locus.

**Supplementary Table 6:** Summary statistics for linear regression HLA interaction model with T2D status adjusted for age, age^2, sex, race, PC1-10 and rs9274619:A, where the outcome was normalized log(TG). The correlation coefficient of each HLA allele to rs9274619:A is documented. The HLA alleles are ordered based on their significance, out of the total 362 HLA alleles 331 alleles had interaction summary statistics.

PC – Principal Components; T2D – Type 2 Diabetes; TG – Triglycerides.

**Supplementary Table 7:** Summary statistics for logistic regression model for PheWAS where disease conditions used as outcomes, adjusted for age, age^2, sex, race and PC1-10. The effects from the rs9274619:A interacting with T2D status is documented and the disease conditions are ordered based on significance.

PC – Principal Components; PheWAS – Phenome Wide Association Studies; T2D – Type 2 Diabetes.

**Supplementary Table 8:** The 45 significant disease conditions identified using interaction model were analyzed using logistic regression, adjusted for age, age^2, sex, race and PC1-10 and stratified by T2D status. The summary statistics from T2D and non-T2D models are tabulated, where the phenotypes are ordered based on T2D beta.

PC – Principal Components; T2D – Type 2 Diabetes.

**Supplementary Table 9:** Multiple linear main and interaction models used to validate the influence of T1D and LADA samples.

LADA – Latent Autoimmune Diabetes in Adults; T1D – Type 1 Diabetes.

**Supplementary Table 10:** Interaction model between the four LADA loci and T2D with TG as outcome

LADA – Latent Autoimmune Diabetes in Adults; T2D – Type 2 Diabetes; TG – Triglycerides.

**Supplementary Table 11:** Interaction p-values between the four LADA loci and T2D with T1D related and hyperlipidemia related phenotypes as outcomes. The top 5 and bottom 5 phenotypes significantly associated with rs9274619:A were selected (Fig. 3). All four loci have significant interaction with diabetes related diseases, but not with obesity related phenotypes.

LADA – Latent Autoimmune Diabetes in Adults; T1D – Type 1 Diabetes; T2D – Type 2 Diabetes.

**Supplementary Table 12:** Summary statistics for linear regression main model with fat, diet and biomarkers phenotypes as outcomes, adjusted for age, age^2, sex, race and PC1-10. The phenotypes are orders based on p-value.

PC – Principal Components.

**Supplementary Table 13:** Summary statistics for linear interaction model with fat, diet and biomarkers phenotypes as outcomes, adjusted for age, age^2, sex, race and PC1-10. The phenotypes are orders based on p-value.

PC – Principal Components.

**Supplementary Table 14:** Summary statistics for linear regression main model with metabolomic phenotype as outcomes, adjusted for age, age^2, sex, race and PC1-10. The effects from rs9274619:A are documented and the metabolomes are ordered based on significance.

PC – Principal Components.

**Supplementary Table 15:** Summary statistics for linear regression interaction model with metabolomic phenotype as outcomes, adjusted for age, age^2, sex, race and PC1-10. The effects from rs9274619:A interacting with T2D status is documented and the metabolomes are ordered based on significance.

PC – Principal Components; T2D – Type 2 Diabetes.

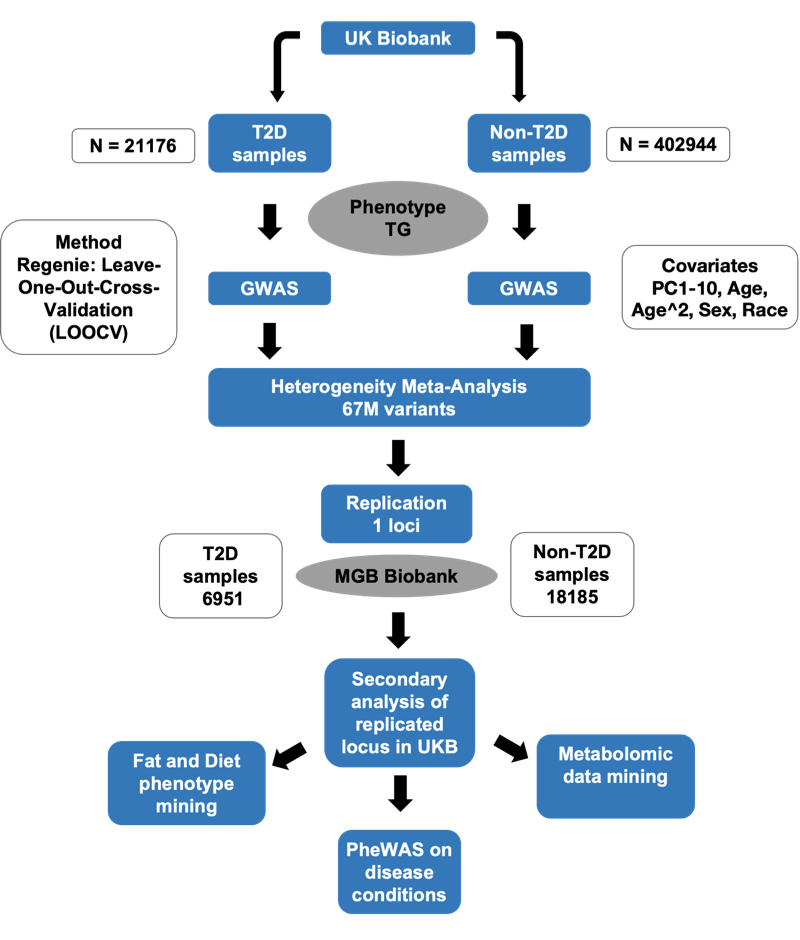

**Supplementary Figure 1: Overall study schematic.**

We carried out stratified GWAS on UKB discovery cohort based on T2 status, using Regenie LOOCV models adjusting for age, age^2^, sex, race, and PC1-10. We implemented heterogeneity analysis to identify loci that was differentially associated between the two strata. Out of the 67M variants analyzed, only one locus achieved genome-wide significance. We replicated the significant locus using MGBB, an independent cohort, and further analyzed the lead variant using various secondary analysis in the discovery cohort.

GWAS – Genome wide association; LOOCV - leave-one-out-cross-validation; MGBB – Mass General Brigham Biobank; T2D – Type 2 Diabetes; UKB – UK Biobank.

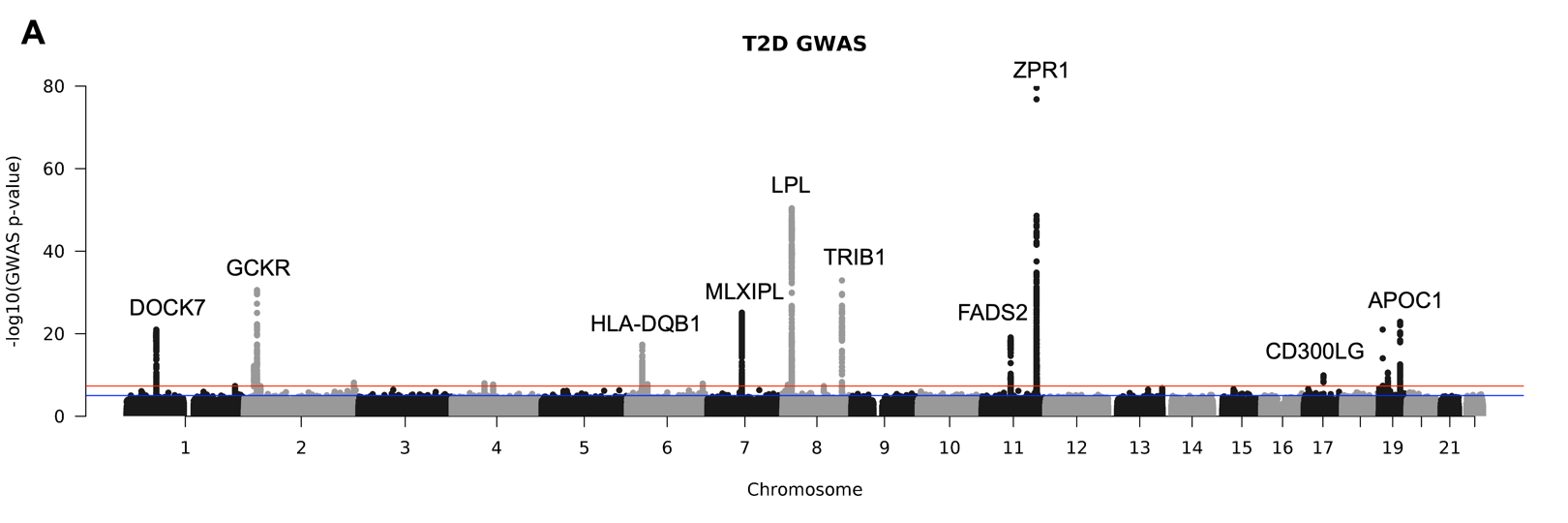

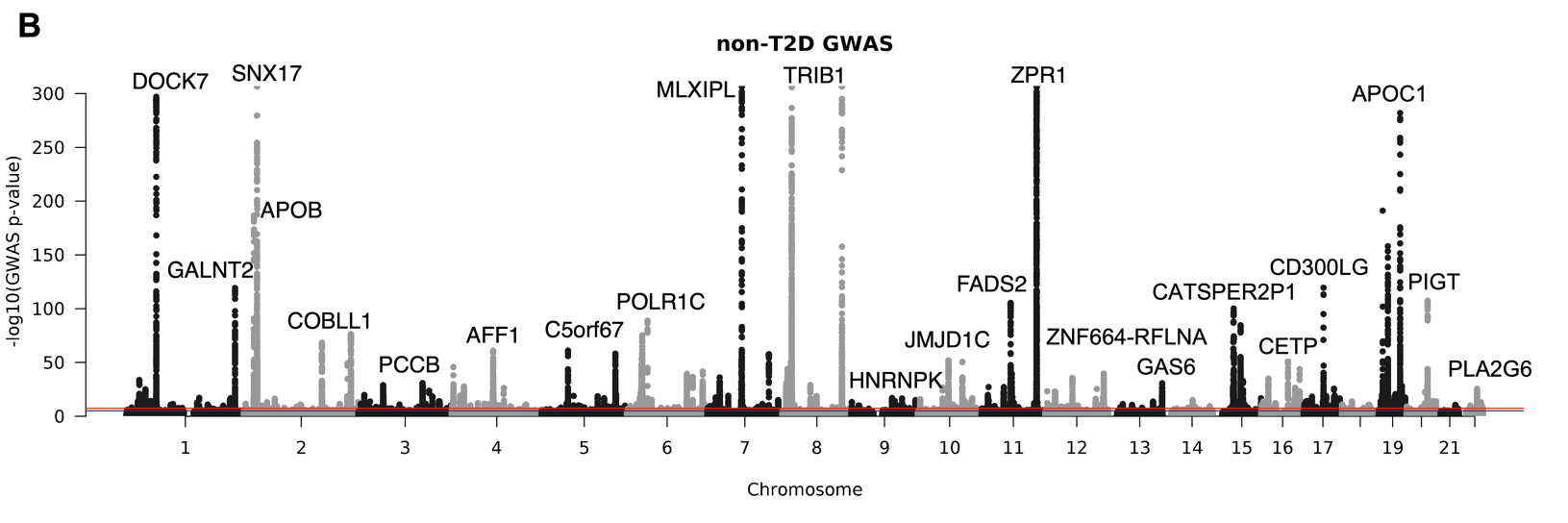

**Supplementary Figure 2: Manhattan (MH) plots for T2D stratified GWAS.**

A) MH plots for T2D GWAS B) MH plots for non-T2D GWAS.

Genes near to the most significant lead variant in each loci are documented, full list of lead SNPs are tabulated in supplementary table 1. Red line: Genome significance (p-value=5x10^-8^), Blue line: Suggestive significance (p-value=1x10^-5^).

GWAS – Genome wide association studies; MH – Manhattan; T2D – Type 2 Diabetes

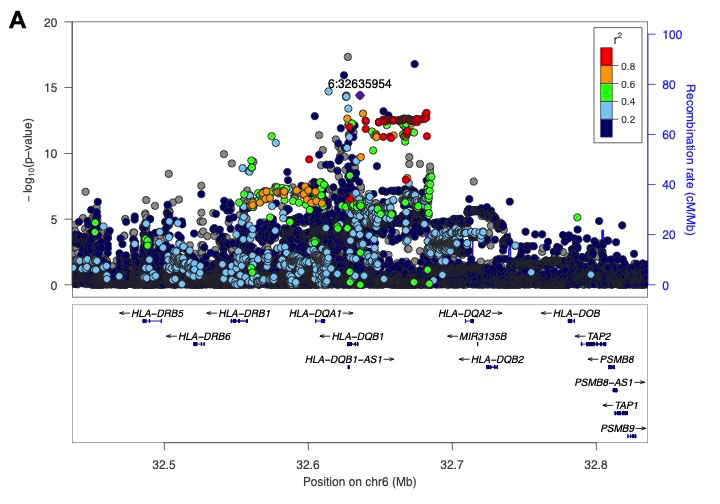

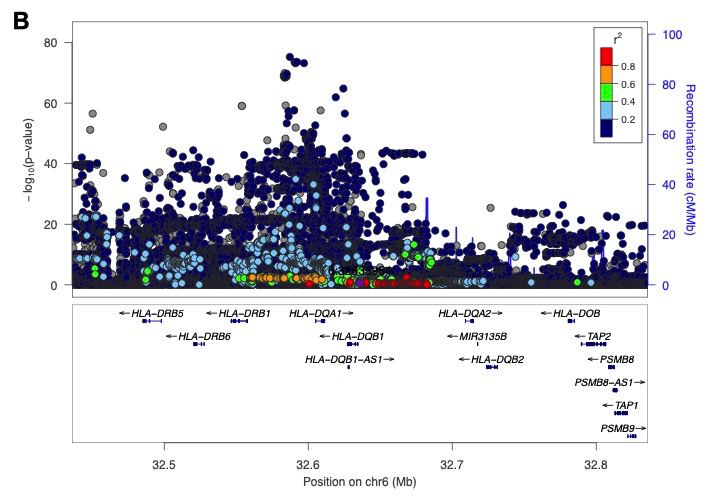

**Supplementary Figure 3: *HLA-DQB1* lead variant locus in T2D and non-T2D strata.**

A) Locus zoom plot for *HLA-DQB1* loci in T2D strata. B) Locus zoom plot for *HLA-DQB1* loci in non-T2D strata.

X-axis defines the genomic position where variants +/-500 kb on either side of rs9274619 - chr6:32635954:G:A (grc37) is mapped on the genome. The variants are colored based on the r^2^ with the lead variant and the genes are mapped based on their genomic position. Y-axis is the -log10(p-values) from the respective strata and the scale of y-axis is different between the two plots.

HLA – Human Leukocyte Antigen; T2D – Type 2 Diabetes

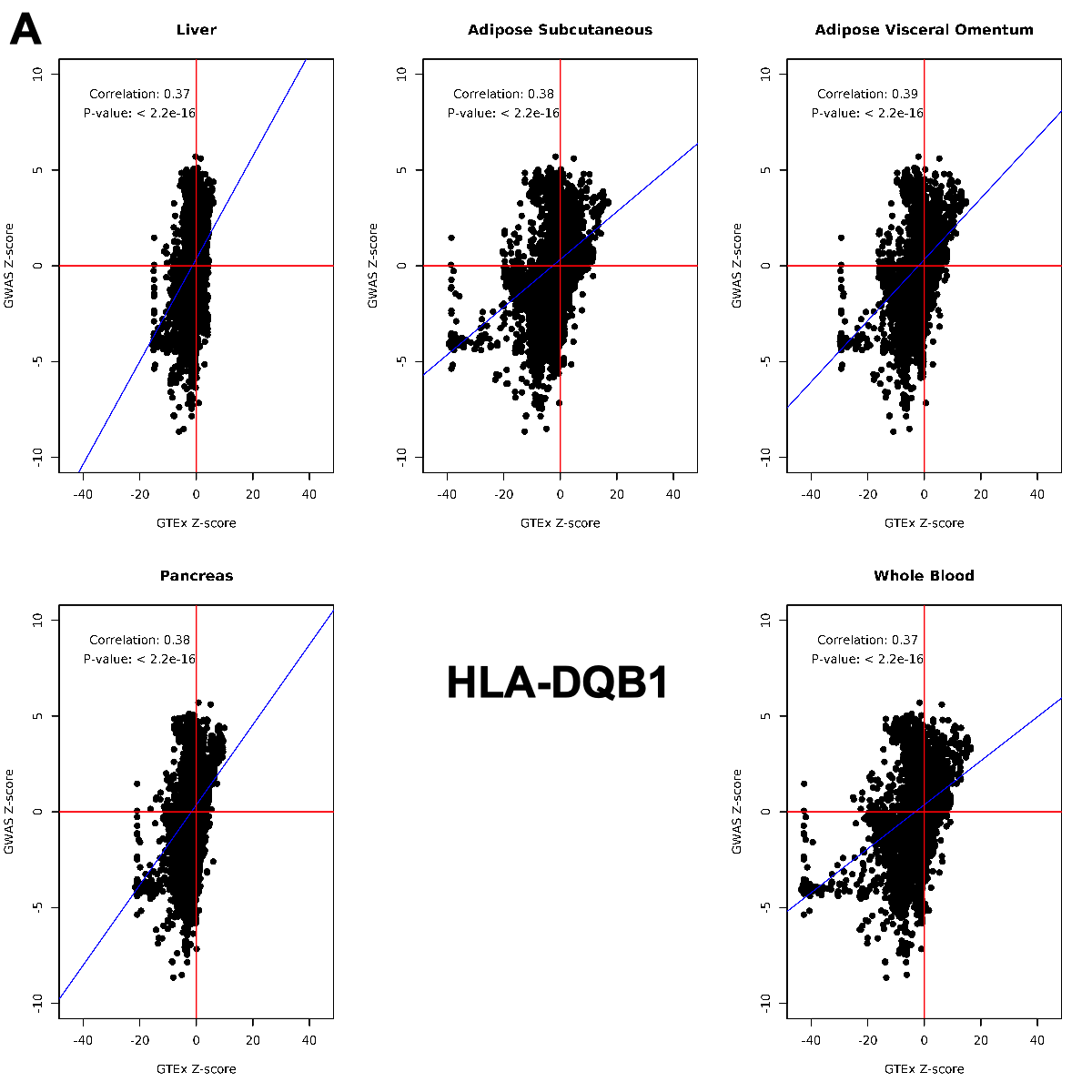

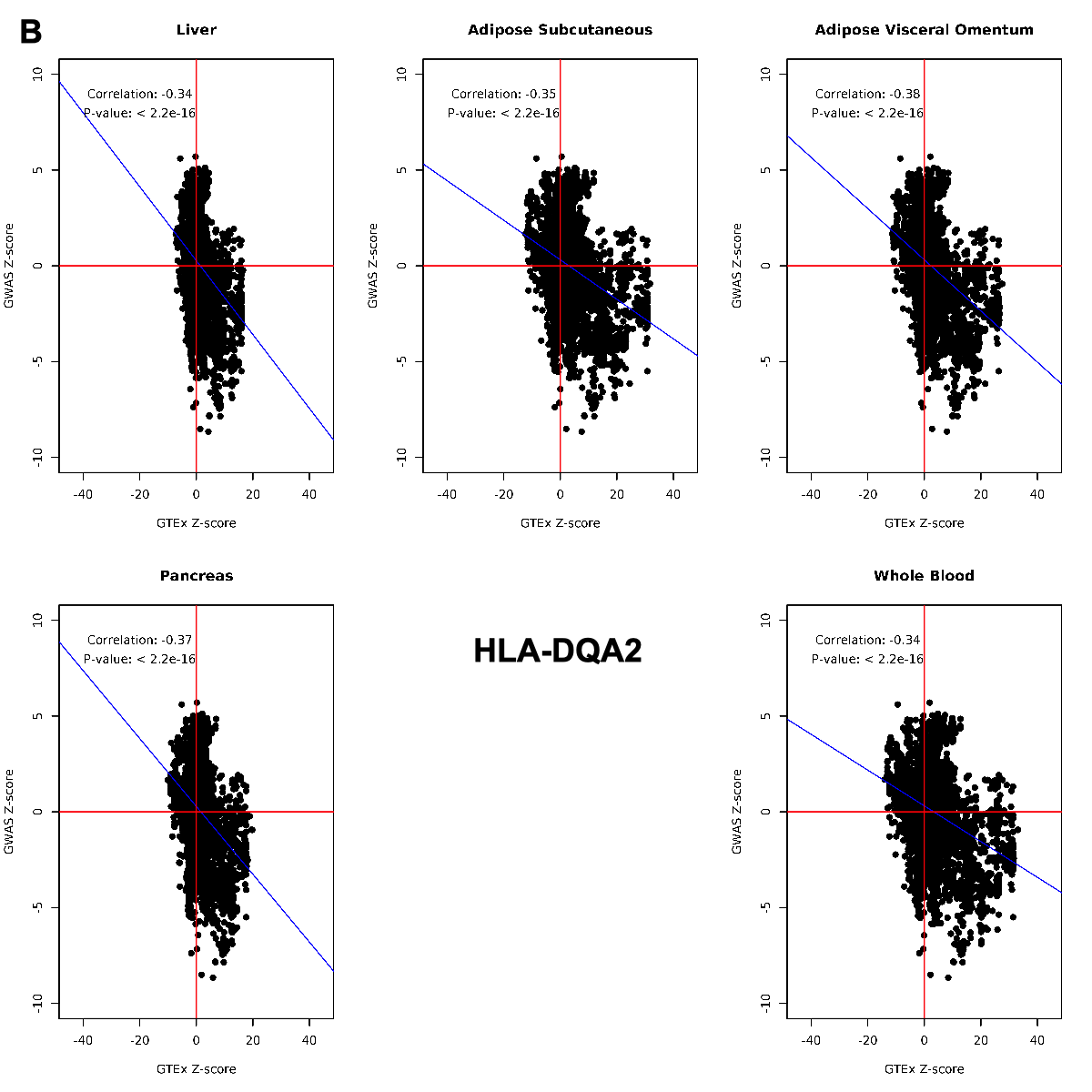

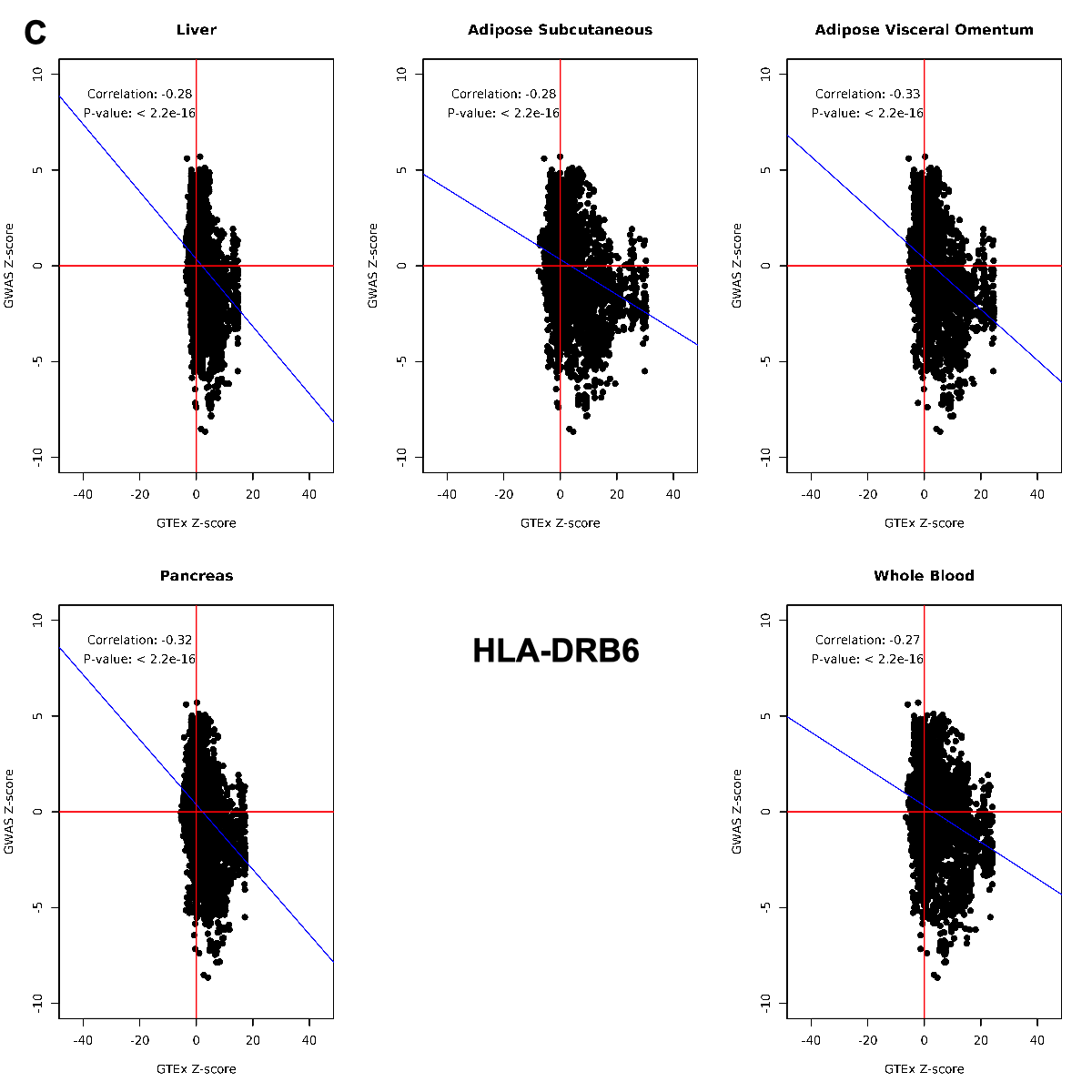

**Supplementary Figure 4: Correlation between GTEx and GWAS Z-scores of eQTLs from *HLA-DQB1*, *HLA-DQA2* and *HLA-DRB6*.**

Z-scores were calculated from T2D GWAS and GTEx (version 8) summary statistics for all the eQTLs for the three genes. Pearson correlation coefficient was calculated, and scatter plots were generated for eQTL data from five different tissues. Most of the TG lowering variants increases the expression of *HLA-DQA2/HLA-DRB6*, whereas decreases the expression of *HLA-DQB1*.

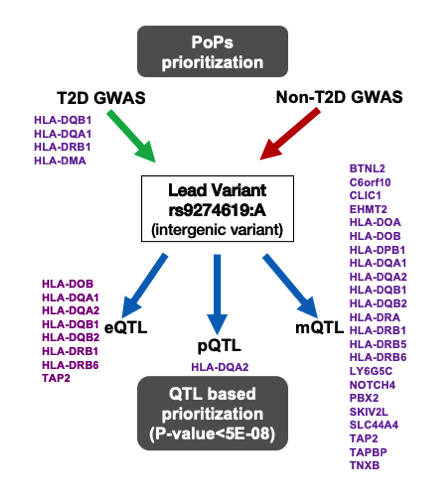

**Supplementary Figure 5: Gene prioritization using PoPS and QTL mining**

PoPS method was used to prioritize genes using GWAS summary statistics from both T2D and non-T2D stratum. Multiple HLA genes were prioritized, where *HLA-DQB1* topped the list. eQTL, pQTL and mQTL curation of rs9274619:A from various public repositories mapped the lead variant to multiple HLA-genes, where *HLA-DQA2* was identified by all three QTL searches.

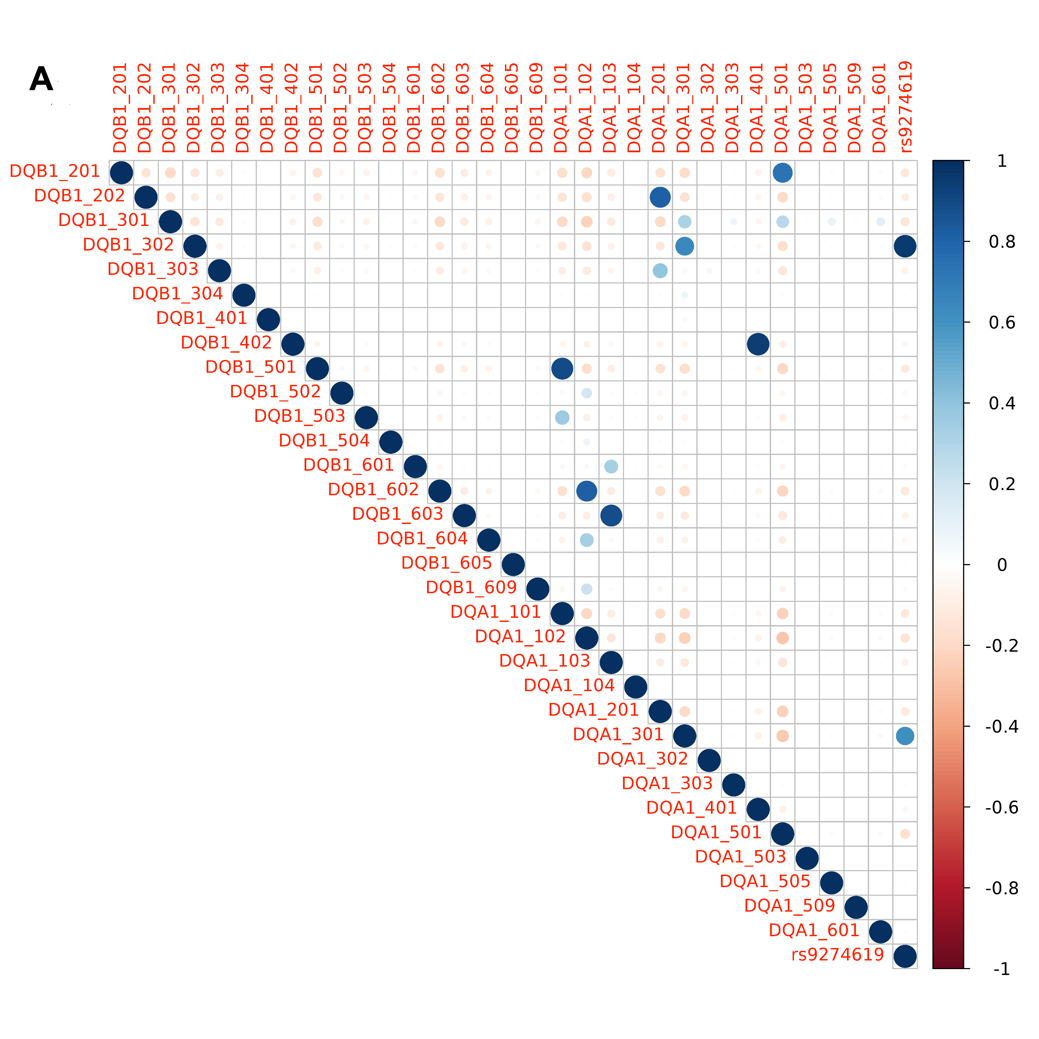

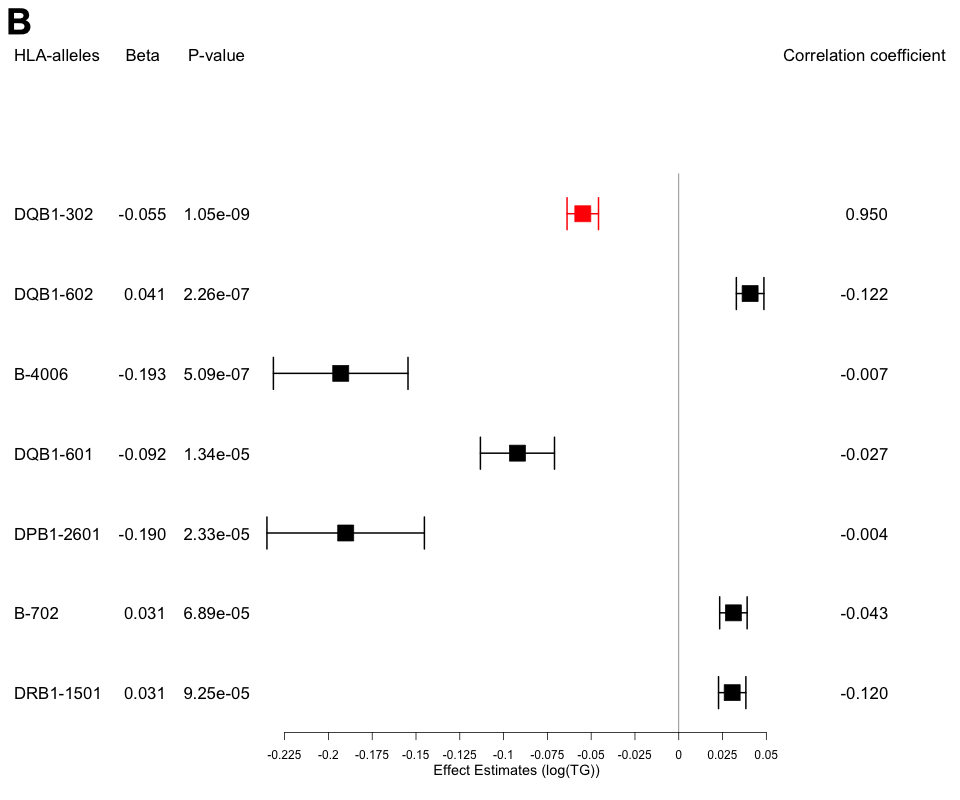

**Supplementary Figure 6: Imputed HLA alleles and its correlation with variant of interest - rs9274619:A.**

A) Correlation between rs9274619:A and HLA alleles in DQB1 and DQA1 class. DQB1-302 is the most strongly correlated allele. B) Forest plot showing the different alleles that passed the Bonferroni correction on interacting with T2D with log(TG) as outcome, the model was adjusted age, age^2^, sex, race, PC1-10 and rs9274619:A. DQB1-302 allele is the only allele with significant interaction with T2D and highly correlated to rs9274619:A (mapped in red).

HLA – Human Leukocyte Antigen; T2D – Type 2 Diabetes; TG – Triglycerides; VOI – Variant of interest.
